# *RPE65*-related retinal dystrophy: mutational and phenotypic spectrum in 45 affected patients

**DOI:** 10.1101/2021.01.19.21249492

**Authors:** R Lopez-Rodriguez, E Lantero, F Blanco-Kelly, A Avila-Fernandez, I Martin Merida, M del Pozo-Valero, I Perea-Romero, O Zurita, B Jiménez-Rolando, ST Swafiri, R Riveiro-Alvarez, MJ Trujillo-Tiebas, E Carreño Salas, B García-Sandoval, M Corton, C Ayuso

## Abstract

**Background:** Biallelic pathogenic *RPE65* variants are related to a spectrum of clinically overlapping inherited retinal dystrophies (IRD). Most affected individuals show a severe progression, with 50% of patients legally blind by 20 years of age. A better knowledge of the mutational spectrum and the phenotype-genotype correlation in *RPE65*-related IRD is needed.

**Methods:** Forty-five affected subjects from 27 unrelated families with a clinical diagnosis of *RPE65*-related IRD were included. Clinical evaluation consisted on self-reported ophthalmological history and objective ophthalmological examination. Patients’ genotype was classified accordingly to variant class (truncating or missense) or to variant location at different protein domains. Main phenotypic outcome was age at onset (AAO) of the symptomatic disease and a Kaplan–Meier analysis of disease symptom event-free survival was performed.

**Results:** Twenty-nine different *RPE65* variants were identified in our cohort, 7 of them novel. Most frequent variants were p.(Ile98Hisfs^*^26), p.(Pro111Ser) and p.(Gly187Glu) accounting for the 24% of the detected alleles. Patients carrying two missense alleles showed a later disease onset than those with 1 or 2 truncating variants (Log Rank test p<0.05). While the 60% of patients carrying a missense/missense genotype presented symptoms before or at the first year of life, almost all patients with at least 1 truncating allele (91%) had an AAO ≤1 year (p<0.05).

**Conclusion:** Our findings suggest an association between the type of the *RPE65* carried variant and the AAO. Thus, our results provide useful data on *RPE65*-associated IRD phenotypes which may help to improve clinical and therapeutic management of these patients.

## INTRODUCTION

Leber congenital amaurosis (LCA; MIM #204000) is the earliest and most severe form of inherited retinal dystrophies (IRD), with a prevalence of ≈1 in 50000 individuals worldwide, accounting for the 5% of IRD [1]. LCA is the most frequent cause of inherited blindness in children [2, 3]. Predominantly inherited in an autosomal recessive (AR) manner, LCA appears as severe visual dysfunction from birth or before the first year of life, characterized mainly by sensory nystagmus, markedly diminished or abolished rod and cone electroretinogram (ERG), abnormal pupil response, loss of visual acuity and variable fundus appearance [4, 5]. Currently, 25 genes have been implicated in the etiopathogenesis of LCA (*RetNet, last accession* on December 2020, https://sph.uth.edu/retnet/) playing a wide variety of critical roles in the retinal development and function.

The *RPE65* gene, located at Chr1p.31.3 and consisting of 14 exons and a 2.7kb transcript [6], encodes the retinoid isomerase Retinal Pigment Epithelium-specific 65kDa (RPE65) protein. In the visual cycle, this enzyme is crucial for regenerating the active chromophore 11-*cis*-retinal in the RPE through all-*trans*-retinyl ester isomerization [7]. In this context, RPE65 dysfunction results in all-*trans*-retinyl ester accumulation and reduced or absent levels of visual pigment in photoreceptors [8]. The RPE65 protein structure is formed by 7 β-blades (the most conserved domain among carotenoid cleavage oxigenases), supporting the active centre of the molecule, connected by α-helices and loops [9]. RPE65 also displayed an external loop (EL) in residues 84 to 133 that includes the endoplasmic reticulum (ER) membrane binding region (MBR). This structure guarantees the specific and correct orientation of the substrate for the cleavage and isomerization [10–12]. In addition, RPE65 has 4 conserved histidine residues and 3 secondary glutamine residues that stabilize and coordinate an iron cofactor required for catalysis.

Biallelic pathogenic *RPE65* variants have been related to a spectrum of clinically overlapping IRD ranging in severity and age of onset from LCA, early-onset retinitis pigmentosa (EORP) and early-onset cone-rod dystrophies [13]. *RPE65* variants represent approximately 6 to 16% and 2% of LCA and EORP cases, respectively [14– 16]. So far, more than 200 different *RPE65* variants have been identified in public databases [17]. Despite of most affected individuals showing a severe progression, with a 50% of patients legally blind by age 20 years, a clear correlation between phenotypic severity and genotype has not been established yet [1, 18, 19].

Genetic diagnosis of *RPE65*-associated retinopathies is currently essential for treatment due to the recent approval by the U.S. FDA (Food and Drug Administration) and the European Medicines Agency (EMA) for first gene replacement therapy for IRD [20]. *Luxturna* (voretigene neparvovec-rzyl) is a recombinant adeno-associated virus serotype 2 (AAV2) vector that delivers a functional copy of *RPE65* into viable RPE cells through a single subretinal injection. This treatment improves ophthalmologic symptoms and reduces vision loss [21]. Given the substantial loss of vision in patients with *RPE65* biallelic mutations during the first three decades of life, timely genetic characterization becomes crucial to the success of gene therapies. Thus, the better knowledge of specific phenotype-genotype correlations in terms of progression and severity of the *RPE65*-associated IRD will definitively help to optimize this new therapy, whose ultimate goal is to preserve functional RPE and photoreceptor cells.

Therefore, our aim is to study genotype-phenotype correlations in 45 IRD patients from 27 families that have been genetically and clinically characterized with biallelic *RPE65* mutations.

## PATIENTS AND METHODS

### Subjects and clinical evaluation

Patients were ascertained from the IRD database of the Fundación Jiménez D^í^az (FJD) University Hospital which contains almost 30 years of collected data. To date, the FJD-IRD database includes 1042 non-syndromic IRD families genetically characterized from a total cohort of 2083 families with autosomal recessive (AR) or sporadic patterns [22]. A total of 45 affected subjects from 27 unrelated families carrying biallelic causative variants in *RPE65* were included in this study. Seven families from our *RPE65* cohort have been previously included in different studies [23–25]. All subjects or their legal guardians provided written informed consent prior to their inclusion in the study. The Research Ethics Committee of our hospital approved the study protocol that adhered the tenets of the Declaration of Helsinki and subsequent reviews.

*RPE65*-associated patients were referred with a clinical diagnosis of LCA, EORP or RP, defined as previously described [26]. Clinical evaluation consisted of a self-reported ophthalmological history, recorded from questionnaires that include the age at onset (AAO) of main symptoms, i.e, visual acuity (VA) loss, visual field (VF) reduction and night blindness (NB), as well as the age at diagnosis and family history. Objective ophthalmological examination including full-field electroretinography (ERG), funduscopy, VF testing, best-corrected VA (BCVA) tests measurements in decimal scale, and in some cases, optical coherence tomography (OCT) and fundus autofluorescence (FAF), have been also performed.

### Genetic analysis

Genomic DNA of probands and their relatives was isolated from EDTA-collected peripheral blood samples using an automated DNA extractor (*MagNAPure Compact system, Roche Applied Science, Penzberg, Germany*). Different strategies were used to perform molecular screening of *RPE65* over time. First, commercial APEX-based genotyping microarrays (LCA or ARRP chips, Asper Biotech, Tartu, Estonia) were used to analyzed 116 known pathogenic *RPE65* variants [25]. Further Sanger sequencing of all exons was carried out to in those patients carrying one *RPE65* allele. PCR primers are listed in Supplementary Table 1. Uncharacterized patients were further analyzed using targeted custom Next Generation Sequencing (NGS) panels based on Haloplex (*Agilent Technologies, Santa Clara, USA*) [27] or Molecular Inversion Probes (*MIPS*, [28]). More recently, commercial clinical exome approaches were routinely performed using *TruSight One Sequencing panel kit (Illumina, San Diego, USA) or Clinical Exome Solution (Sophia Genetics, Boston, USA*) [29]. All NGS libraries were obtained following manufacturer instructions and sequenced on Miseq or NextSeq500 Illumina systems. All SNVs (Single Nucleotide Variations) and CNVs (Copy Number Variations) identified in the index patient were further confirmed and segregated in families, whenever possible (n=21 families), by Sanger sequencing or multiplex ligation-dependent probe amplification (*MLPA, SALSA P221 LCA mix-1, MRC, Holland*), respectively.

### Pathogenicity assessment of *RPE65* variants

All *RPE65* variants were named according to the reference sequence NM_000329.3 and classified following the recommendations of the American College of Medical Genetics and Genomics (ACMG) [30]. Genetic variants were considered pathogenic according to: 1) their allele frequency in GnomAD (*http://gnomad.broadinstitute.org/*), 2) disease causing *RPE65* variants previously reported in literature, the Human Gene Mutation (HGMD, *Stenson PD et al. 2017*) or *ClinVar* databases (*https://www.ncbi.nlm.nih.gov/clinvar/*); 3) family segregation when DNA from other relatives was available; 4) pathogenic *in silico* prediction. Missense variants were predicted as damaging by at least 3 out of several predictor tools (*SIFT, Polyphen-2, Mutation Taster, M-CAP, CADD*, among others). Canonical and noncanonical splicing variants were assessed using 5 predictors (*MaxEntScan, Human Splicing Finder, Splice Site Finder-like, NNSPLICE and GeneSplicer*) using the *Alamut* software (*Interactive Biosoftware, Rouen, France*).

Pathogenic variants were classified accordingly to their functional impact as truncating (nonsense, frameshift, splicing variants and multiexon deletion) or missense. Also, variants were grouped based on their location at the putative functional and structural RPE65 protein domains previously described [10–12].

### Haplotype analysis

Haplotypes of the *RPE65* locus in families carrying same *RPE65* variants were analyzed by genotyping 6 microsatellite markers (D1S2803, D1S2806, D1S368, D1S2829, D1S448 and D1S1162) flanking the *RPE65* gene in order to determine possible founder effects. Primers and the relative position of each marker referred to *RPE65* (Chr1[hg19]) are detailed in Supplementary Table 2.

### Genotype-phenotype correlation

Genotypes were stratified depending on: i) the functional effect of the *RPE65* variants (patients with 0, 1 or 2 truncating alleles) or ii) the variant location at EL (84 to 133 residues) of the RPE65 protein (0, 1 or 2 alleles at the EL). Main phenotypic outcomes to analyze were the AAO of the symptomatic visual loss, self-reported by the patient or tenant, and/or the age at diagnosis. The AAO of the main characteristic disease symptoms (VA loss, VF reduction and NB) were also analyzed.

Statistical association between genotype subgroups and phenotypic features was assessed by Kaplan–Meier analysis of ophthalmological symptoms event-free survival (long rank test) and Chi-squared test. Two-tailed *p-values* below 0.05 were considered statistically significant.

## RESULTS

### Mutational spectrum

A total of 27 families were characterized with biallelic disease-causing *RPE65* variants, leading a prevalence that accounts for 2.6% (CI 95%= 1.6-3.5%) of the 1042 non syndromic IRD genetically characterized families with AR or sporadic pattern in our centre. Fifteen of these 27 families were initially catalogued as sporadic cases and 12 as AR cases accounting for a total of 45 affected cases. Pathogenic *RPE65* variants were found in homozygosis in 9 families (Table 1). Pedigrees of the 27 families are available upon request to the corresponding author.

Twenty-nine different *RPE65* variants have been identified in our cohort, 7 of them were novel and other 7 were previously described for the first time in our cohort (Figure 1A and Table 2). All novel missense variants were classified as damaging or probably pathogenic by different *in silico* predictors (Supplementary Table 3). Most of the detected variants were located in coding regions (n=25) and the EL was the structural protein domain most frequently mutated, accounting 5 different variants located at this domain for the 28% of the total of alleles detected in probands (Figure 1B and Table 2). Three variants were found at intronic regions modifying splice sites and a complete gene deletion was also found. Missense variants were the most frequently found (n=17, 58.6%), followed by frameshift (n=4, 13.8%), nonsense (n=4, 13.8%), splicing variants (n=3, 10.3%) and finally the complete *RPE65* deletion (n=1; 3.4%), as described in Figure 1C.

**Figure 1.**
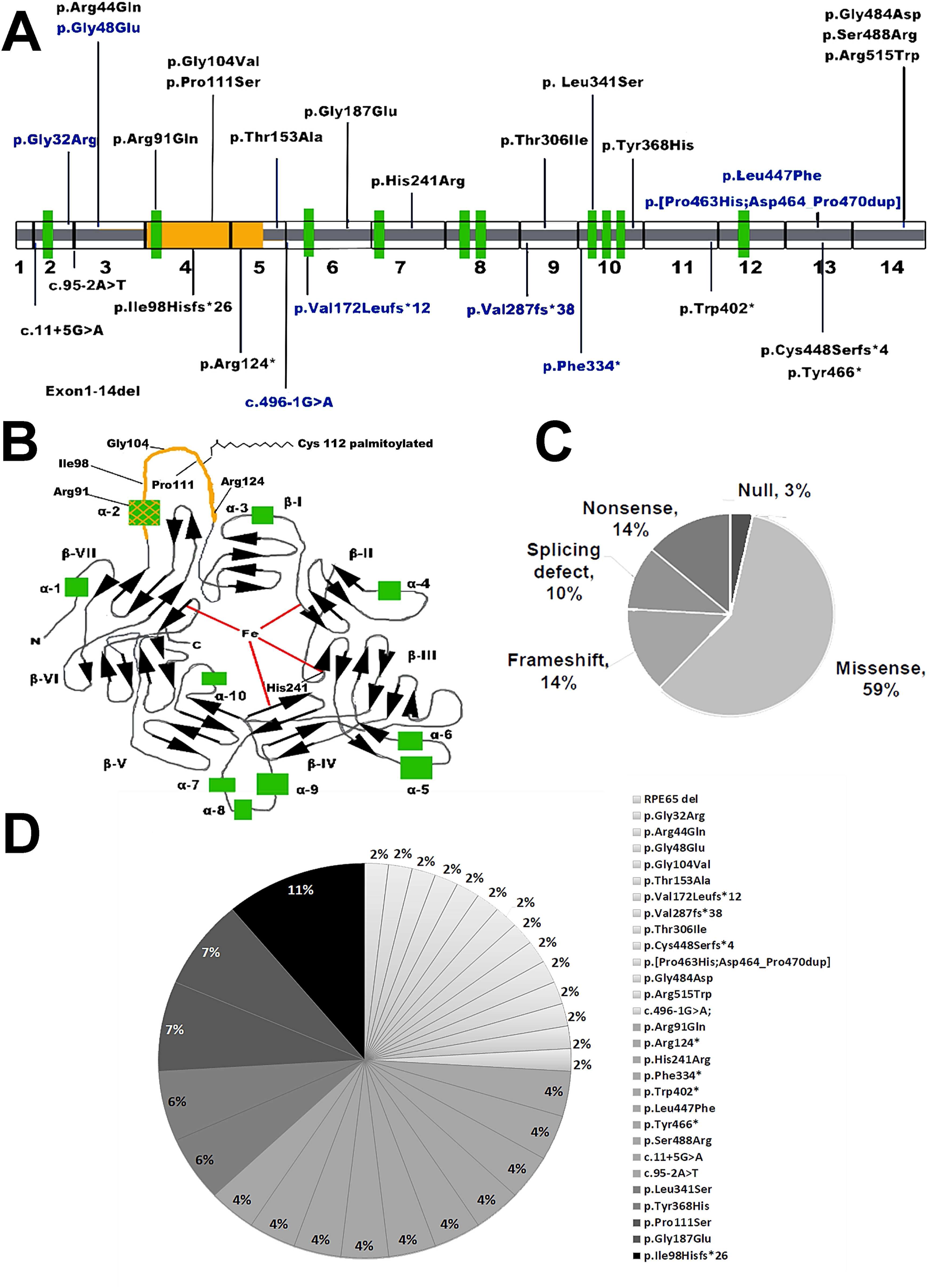
Schematic representation of the gene and protein structure of *RPE65* and the spectrum of all pathogenic variants found in the FJD IRD cohort. *A)* Representation of the 14 exons of RPE65. Pathogenic variants detected are depicted over the gene. Novel variants detected firstly in our cohort are in blue. B) Representation of the RPE65 protein adapted from Kiser et al. (2009) [12]. Structural protein domains are depicted along the gene: black arrows and grey lines indicate the 7 bladed beta propeller domain (β-I to VII), green squares indicate α-helix structure, the external loop (EL) is depicted in yellow and red lines indicated residues for iron coordination (His180, His241, His313 and His527). C) Frequency of RPE65 variants accordingly to their functional impact. D) Allelic frequency of each of the 29 variants detected in the 27 probands from our RPE65 cohort.

**Table 1.** Genotypes of the 27 families with biallelic disease-causing *RPE65* variants identified at the FJD IRD cohort.

| Family_ID | Allele 1 |  |  | Allele 2 |  |  | Zygosity | Patients (n=45) |
| --- | --- | --- | --- | --- | --- | --- | --- | --- |
|  | Ex/Int | Nucleotide | Aminoacid | Ex/Int | Nucleotide | Aminoacid |  |  |
| RPE65_1 | IVS 1 | c.11+5G>A | p.? | 4 | c.331C>T | p.Pro111Ser | HT Comp | 1 |
| RPE65_2* | IVS 1 | c.11+5G>A | p.? | 13 | c.1341_1342 dup | p.Cys448Serfs*4 | HT Comp | 1 |
| RPE65_3 | 2 | c.94G>C | p.Gly32Arg | 6 | c.560G>A | p.Gly187Glu | HT Comp<sup>a</sup> | 1 |
| RPE65_4<sup>#</sup> | IVS 2 | c.95-2A>T | p.? | 4 | c.311G>T | p.Gly1104Val | HT Comp | 1 |
| RPE65_5<sup>&</sup> | IVS 2 | c.95-2A>T | p.? | 5 | c.457A>G | p.Thr153Ala | HT Comp | 2 |
| RPE65_6 | 3 | c.131G>A | p.Arg44Gln | 6 | c.514_515del | p.Val172Leufs*12 | HT Comp<sup>a</sup> | 4 |
| RPE65_7 | 3 | c.131G>A | p.Arg44Gln | 9 | c.859delG | p.Val287fs*38 | HT Comp | 3 |
| RPE65_8 | 4 | c.272G>A | p.Arg91Gln | 4 | c.272G>A | p.Arg91Gln | Homo<sup>a</sup> | 3 |
| RPE65_9 | 4 | c.292_311del | p.Ile98Hisfs*26 | 4 | c.292_311del | p.Ile98Hisfs*26 | Homo | 3 |
| RPE65_10<sup>#</sup> | 4 | c.292_311del | p.Ile98Hisfs*26 | 13 | c.[1388C>A; 1390_1410dup] | p.[Pro463His; Asp464_Pro470dup] | HT Comp | 1 |
| RPE65_11 | 4 | c.292_311del | p.Ile98Hisfs*26 | 10 | c.1102T>C | p.Tyr368His | HT Comp | 2 |
| RPE65_12 | 4 | c.331C>T | p.Pro111Ser | 4 | c.331C>T | p.Pro111Ser | Homo | 2 |
| RPE65_13<sup>#</sup> | 4 | c.331C>T | p.Pro111Ser | 14 | c.1451G>A | p.Gly484Asp | HT Comp | 1 |
| RPE65_14<sup>#</sup> | 5 | c.370C>T | p.Arg124* | 1-14 | Gene deletion of at least exon 1 to 14 |  | HT Comp | 1 |
| RPE65_15 | 5 | c.370C>T | p.Arg124* | 10 | c.1001_1002del | p.Phe334* | HT Comp<sup>a</sup> | 2 |
| RPE65_16 | IVS 5 | c.496-1G>A | p.? | 10 | c.1022T>C | p. Leu341Ser | HT Comp | 1 |
| RPE65_17 | 6 | c.560G>A | p.Gly187Glu | 6 | c.560G>A | p.Gly187Glu | Homo | 4 |
| RPE65_18 | 6 | c.560G>A | p.Gly187Glu | 10 | c.1001_1002del | p.Phe334* | HT Comp | 1 |
| RPE65_19 | 7 | c.722A>G | p.His241Arg | 7 | c.722A>G | p.His241Arg | Homo | 2 |
| RPE65_20 | 9 | c.917C>T | p.Thr306Ile | 10 | c.1022T>C | p.Leu341Ser | HT Comp | 1 |
| RPE65_21 | 10 | c.1022T>C | p.Leu341Ser | 14 | c.1543C>T | p.Arg515Trp | HT Comp<sup>a</sup> | 1 |
| RPE65_22 | 10 | c.1102T>C | p.Tyr368His | 10 | c.1102T>C | p.Tyr368His | Homo<sup>a</sup> | 2 |
| RPE65_23 | 11 | c.1205G>A | p.Trp402* | 11 | c.1205G>A | p.Trp402* | Homo | 1 |
| RPE65_24 | 13 | c.1339C>T | p.Leu447Phe | 13 | c.1339C>T | p.Leu447Phe | Homo | 1 |
| RPE65_25 | 13 | c.1398C>G | p.Tyr466* | 14 | c.1464T>A | p.Ser488Arg | HT Comp | 1 |
| RPE65_26* | 13 | c.1398C>G | p.Tyr466* | 14 | c.1464T>A | p.Ser488Arg | HT Comp | 1 |
| RPE65_27 | 4 | c.292_311del | p.Ile98Hisfs*26 | 4 | c.292_311del | p.Ile98Hisfs*26 | Homo<sup>a</sup> | 1 |

ID: family number identification; Ex, exon; Int, intron; Homo, homozygosis; HT Comp, compound heterozygosis.
\*Subjects previously described by *Kumaran et al., 2018* [23] (RPE65\_2 and \_26 is equal to NM\_304 and NM\_313, respectively). <sup>#</sup>subjects previously described by *Kumaran et al., 2020* [24] (RPE65\_4, \_10, \_13 and \_14 is equal to NM\_434, NM\_419, NM\_413 and NM\_423, respectively). <sup>&</sup>Family previously described by *Ávila-Fernández A et al.; 2010* [25] (RPE65\_5 is equal to RP-1115). <sup>a</sup>Families without available samples for segregation study in one or more members of the family.

**Table 2.**
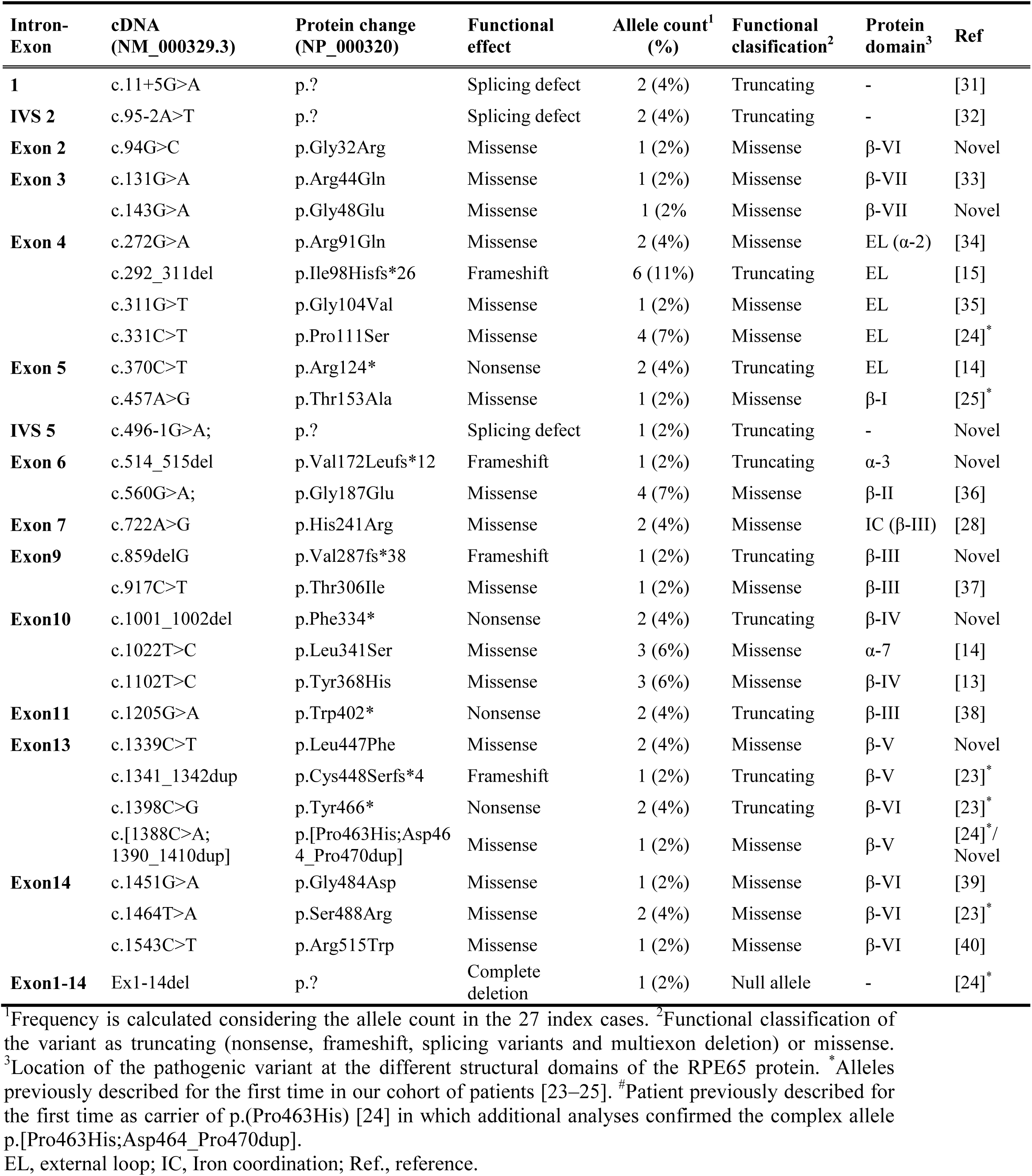
Pathogenic alleles found in our *RPE65* cohort.

The most frequent pathogenic variants in our cohort are p.(Ile98Hisfs^*^26) accounting for the 11%, followed by p.(Pro111Ser) and p.(Gly187Glu) representing each of them the 7% of the total of detected alleles (Figure 1D and Table 2). All families carrying any of these frequent *RPE65* alleles were not related and had different geographic origin; except for 2 families carrying the p.(Gly187Glu) variant (RPE65_17 homozygous and RPE65_18 maternal allele). Haplotype analyses were performed in 6 families carrying p.(Ile98Hisfs^*^26) or p.(Pro111Ser) variants. The p.(Ile98Hisfs^*^26) variant was found in homozygosis in RPE65_9 and RPE65_27 families and in compound heterozygosis in other 2 families (RPE65_10 and RPE65_11). A common haplotype was found for the 3 families with DNA available for this study (RPE65_9, _10 and _11), showing the same number of repeats at D1S2803, D1S448 and D1S1162 markers. In addition, p.(Pro111Ser) was found in 1 family in homozygosis (RPE65_12) and in 2 families in compound heterozygosis (RPE65_1 and RPE65_13). A common haplotype was found for these 3 families showing the same number of repeats at the D1S2803 and D1S448 markers. Haplotype study of p.(Gly187Glu) variant was not carried out due to unavailability of DNA samples.

Interestingly, 2 families (RPE65_25 and RPE65_26) from the same isolated geographical area and without an evident connection between them, carried the same 2 pathogenic alleles in compound heterozygosis (p.(Tyr466^*^) and p.(Ser488Arg)). Analysis of both probands revealed similar haplotypes, despite not being possible to infer the allelic phase of the three markers (D1S2829, D1S448 and D1S1162); thus, possible founder effect cannot be discarded.

### Phenotypic characteristics

Clinical data were available in 35 out of 45 affected subjects (Table 3 and Supplementary Table 4). The most frequent *a priori* diagnosis in probands was LCA (n=18), accounting the *RPE65*-associated LCA the 16.5% of the total of LCA families genetically characterized in our cohort (n=109). Accordingly, most of patients were diagnosed in infancy during the first year of life (Table 3 and Figure 2A). Apart from congenital nystagmus, the earliest clinical sign of visual impairment was NB, present in 32 of the 45 affected cases mostly before or at first year of life (Figure 2A). Nystagmus was present in 16 of 45 patients (Table 3).

**Table 3.** Summary of clinical data in 45 affected subjects.

| Family ID | Patient | Dx | AAO of DS | Nyst (AAO) | NB (AAO) | VF red. (AAO) | VA loss (AAO) | Fundus |
| --- | --- | --- | --- | --- | --- | --- | --- | --- |
| RPE65_1 | P | LCA/CRD | <1 | N | Y (<1) | ≤5 | ≤5 | Possible vascular thinning and peripheral RPE changes in RE |
| RPE65_2 | P | LCA | ≤1 | Y | Y (≤1) | ≤5 | ≤5 | Retinal dystrophy, attenuated vessels |
| RPE65_3 | P | RP | U | U | U | U | U | U |
| RPE65_4 | P | LCA | <1 | Y | U | U | U | U |
| RPE65_5 | P | EORP | ≤1 | N | Y (≤10) | ≤20 | U | Retinal degeneration without spicules or pigment. |
|  | S | U | ≤1 | N | Y (≤1) | ≤1 | 20s | U |
| RPE65_6 | P | LCA/CRD | <20* | Y | Y (<1) | <1 | <1 | Diffuse RPE changes in BE |
|  | S | U | <1 | Y | Y (<1) | <1 | <1 | White retina, bilateral macular affectation, narrow and vascular thinning |
|  | S | U | <1 | N | Y (<1) | <1 | <1 | U |
|  | S | U | U | U | U | U | U | U |
| RPE65_7 | P | LCA | <1 | Y (≤1) | Y (≤1) | ≤1 | ≤1 | Diffuse retinal pigmentation, vascular thinning and pale disc in BE |
|  | S | U | U | U | U | U | U | Diffuse retinal pigmentation, vascular thinning and pale disc in BE |
|  | S | U | U | U | U | U | U | Diffuse retinal pigmentation, vascular thinning and pale disc in BE |
| RPE65_8 | P | EORP | U | U | U | U | U | U |
|  | S | U | U | U | U | U | U | U |
|  | S | U | U | U | U | U | U | U |
| RPE65_9 | P | LCA | <1 | Y (U) | Y (<1) | <1 | ≤1 | Pigment in spicules 360 degrees and posterior pole and vascular thinning |
|  | S | U | <1 | Y | Y (≤1) | ≤1 | ≤1 | U |
|  | S | U | <1 | Y (U) | Y (≤1) | ≤1 | ≤1 | U |
| **RPE65_10** | P | LCA | <1 | Y (<1) | Y (<1) | ≤1 | <1 | Mottled RPE changes |
| **RPE65_11** | P | LCA | <1 | Y | Y (<1) | ≤1 | ≤5 | Mottled RPE changes |
|  | S | U | <1 | N | Y (<1) | <1 | <1 | U |
| **RPE65_12** | P | RP | ≤1 | N | Y (≤5) | 20s | 20s | Diffuse retinal pigmentation and vascular thinning |
|  | S | U | ≤1 | N | Y (<1) | <1 | <1 | Mild macular atrophy |
| **RPE65_13** | P | LCA | ≤1 | N | Y (≤1) | ≤1 | ≤1 | Salt & pepper pigmentation |
| **RPE65_14** | P | LCA/<br>EORP | <1 | Y | Y (<1) | ≤1 | <1 | Normal |
| **RPE65_15** | P | LCA | <1 | Y | Y (≤1) | ≤1 | ≤1 | RPE changes and pale disc |
|  | S | U | U | U | U | U | U | U |
| **RPE65_16** | P | LCA/<br>EORP | <1 | Y (U) | Y (<1) | ≤1 | <1 | Vascular thinning, pigment in spicules in the RE |
| **RPE65_17** | P | RP | <10 | N | Y (<10) | U | U | Pigment in spicules |
|  | S | U | 20s | N | Y (30s) | 40s | 30s | Oblique papillas, paravascular pigment in spicules |
|  | S | U | ≤5 | Y (U) | Y (≤5) | ≤5 | ≤5 | RPE changes and pale disc in BE |
|  | S | U | 20s | N | Y (20s) | 30s | 30s | Pigment in spicules outside arcades 360 degrees |
| **RPE65_18** | P | RP | ≤5 | N | Y (≤5) | ≤5 | U | Pigment in spicules outside arcades 360 degrees |
| **RPE65_19** | P | RP | ≤5 | N | Y (<1) | ≤10 | ≤10 | Pigment in spicules outside arcades 360 degrees and macular atrophy |
|  | S | RP | U | U | U | U | U | Diffuse retinal pigmentation, vascular thinning and pale disc in BE |
| **RPE65_20** | P | RP | ≤1 | N | Y (≤5) | ≤1 | ≤5 | Salt & pepper pigmentation |
| **RPE65_21** | P | LCA/<br>EORP | <1 | N | Y (<1) | <1 | <1 | *Punctata albescens* appearance |
| **RPE65_22** | P | RP | <1 | Y (<1) | U | U | U | Retinopathy, no pigment. Optic atrophy related to congenital glaucoma |
|  | S | U | U | U | U | U | U | U |
| **RPE65_23** | P | LCA | <1 | N | Y (<1) | ≤5 | ≤1 | U |
| **RPE65_24** | P | LCA | <1 | U | Y (<1) | <1 | <1 | U |
| **RPE65_25** | P | LCA | <1 | Y (<1) | Y (<1) | ≤5 | <1 | Pale disc and hypopigmented fundus |
| **RPE65_26** | P | LCA | <1 | N | Y (<1) | <1 | <1 | RPE changes |
| **RPE65_27** | P | LCA/<br>EORP | <1 | U | U | <1 | <1 | Pigment in spicules, vascular thinning and pale disc in BE |

P, Probandus; S, siblings of the index case; Dx, *a priori* diagnosis; LCA, Leber congenital amaurosis; CRD: cone rod dystrophy; EORP, early onset retinitis pigmentosa; RP, retinitis pigmentosa; AAO, age at onset in years; DS, disease symptoms; Nyst, nystagmus; NB, night blindness; VF, visual field; VA, visual acuity; U, unknown; Y, yes; N, no; RPE, retinal pigment epithelium; BE, both eyes; RE, right eye and LE, left eye. \*Indicates age at Dx, patient refers AAO at infancy.

**Figure 2.**
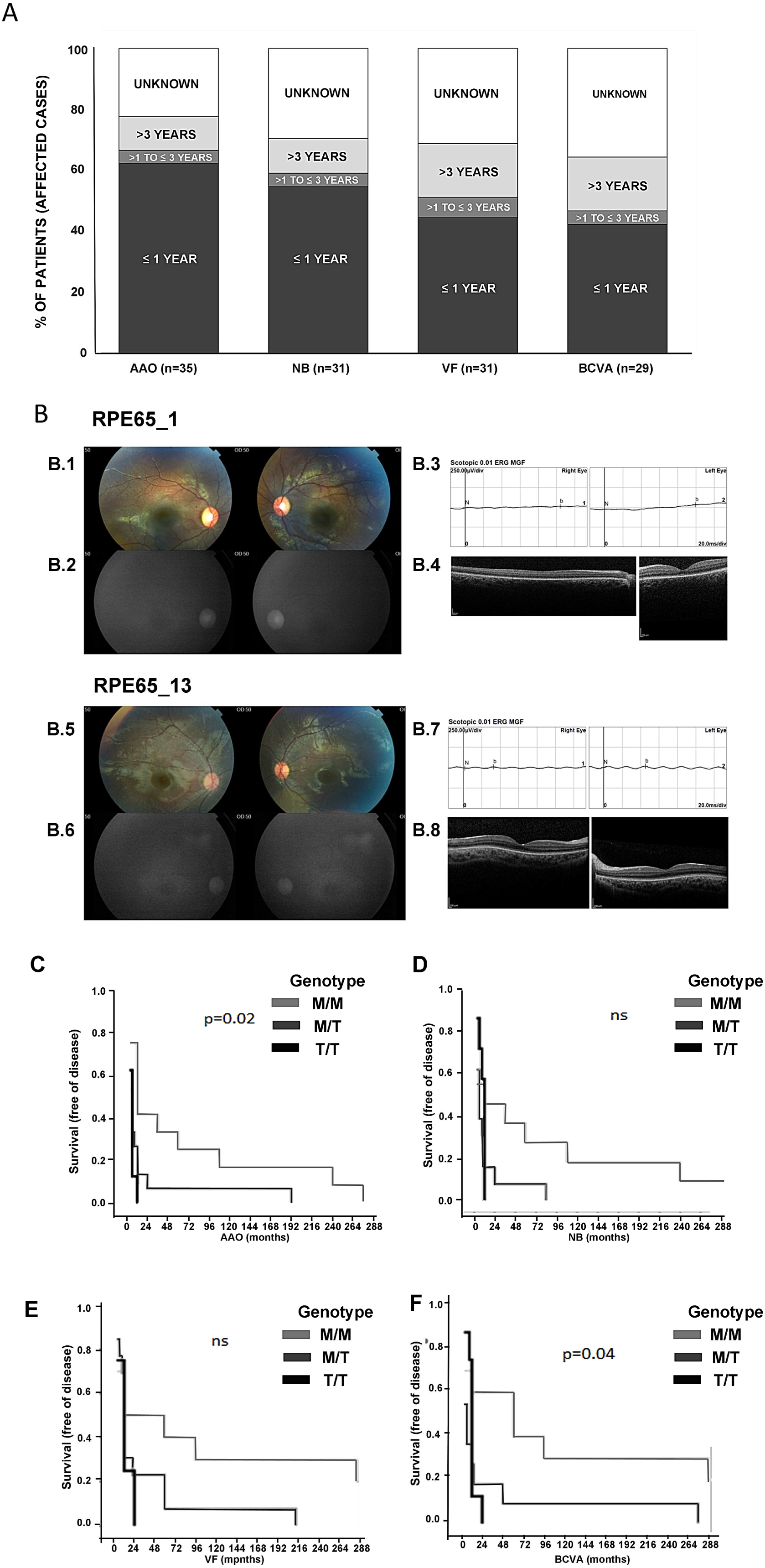
Clinical characteristics and phenotype-genotype correlation: age at onset of disease among patients according to the carried variant class. A) Age at onset (AAO) of the disease and the main signs of visual impairment in the RPE65 cohort. B) Phenotypic features of two patients carrying the novel variant p.Pro111Ser from families RPE65_1 and RPE65_13. Index case RPE65_1: B.1) Colour fundus photographs of both eyes (Left right eye, Right left eye) showing a normal appearance of the retina and optic nerve. B.2) Fundus autofluorescence, showing hyperautofluorescence of the optic nerve, probably related to the relative hypoautofluorescence of the retina with a mild hyperautofluorescent ring perifoveal in both eyes (Left right eye, Right left eye). B.3) Dark adapted 0.01 register of the full-field electroretinogram (ff-ERG) disclosing not registrable signal (Left right eye, Right left eye). B.4) Spectral domain optical coherence tomography (SD-OCT) inferior to fovea in the right eye (left) and through fovea in the left eye (right) showing a mild granularity of the external retina with an apparently preserved ellipsoid zone subfoveally. Index case RPE65_13: B.5) Colour fundus photographs of both eyes (Left right eye, Right left eye) showing a normal appearance of the retina and optic nerve. B.6) Fundus autofluorescence, showing hyperautofluorescence of the optic nerve, probably related to the relative hypoautofluorescence of the retina with a mild hyperautofluorescent ring perifoveal in both eyes (Left right eye, Right left eye). B.7) Dark adapted 0.01 register of the ff-ERG disclosing not registrable signal (Left right eye, Right left eye). B.8) SD-OCT through fovea (Left right eye, Right left eye) showing a mild granularity of the external retina with an apparently preserved ellipsoid zone subfoveally. C) Log Rank test and survival curve (free of disease) of the AAO of the first sign of disease, D) Night blindness, E) Visual field reduction and F) Best-corrected visual acuity loss according to patients’ genotype (0, 1 or 2 truncating alleles). AAO, Age at onset; NB, night blindness; VF, Visual Field; BCVA, Best Corrected Visual Accuity; ns, not significant; M/M, carrier of two missense variants; M/T, carrier of one missense and one truncating allele and T/T, carrier of two truncating alleles

Constricted VF were documented at infancy (<10 years old) in 3 families (RPE65_10, _12 and _14; Supplementary Table 4). Although BCVA was severely reduced in most cases (≤0.1 in 24 of 33 affected subjects with data available), it ranged from hand motion to good visual preservation in 2 siblings from the same family (RPE65_5, compound heterozygous for c.95-2A>T and p.(Thr153Ala)) that displayed BCVA ≥0.7 in both eyes in their adulthood (Supplementary Table 4).

Full field ERG was available in 19 patients showing in all cases an extinguished or altered register (Supplementary Table 4). Fundus changes were not homogeneous among patients and ranged from normal fundus appearance without pigment (RPE65_1 and _12) or isolated areas of retinal atrophy without pigment (RPE65_5 and _20) to retinal dystrophy with attenuated blood vessels (RPE65_2) or pigmented speckles in the whole eye fundus (RPE65_9). In most cases, the retina appeared pale with mild diffuse pigment accumulation. However, no obvious difference in fundus changes appeared to exist among patients carrying different categories of mutations. Only one index case heterozygous for p.(Leu341Ser) and p.(Arg515Trp), showed *fundus albipunctatus*-like changes (RPE65_21, Table 3). Other clinical features were cataracts (n=7), disturbed colour vision (n=8) or congenital glaucoma (n=1, Supplementary Table 4).

Interestingly, p.(Pro111Ser) variant, located close to the key residue Cys112 (Figure 1C), was found in 3 families (RPE65_1, _12 and _13). None of the 4 patients carrying this variant displayed nystagmus and 3 of them showed dyschromatopsia (RPE65_12, 2 siblings and RPE65_13 probandus). Fundus photographs, OCT and ERG records were available for families RPE65_1 and _12 at infancy (≤5 years old, both; Figure 2B). Both patients showed a normal appearance of the retina and optic nerve, hyperautofluorescence of the optic nerve, not registrable ERG signals and a mild granularity of the external retina with an apparently preserved ellipsoid zone subfoveally determined by the OCT (Figure 2B).

In addition, one of the families (RPE65_19) carried a variant in homozygosis (p.(His241Arg)) in one of the crucial residues for iron coordination. Two affected siblings displayed BCVA >0.1 at adulthood but the limited clinical data available in this family makes it difficult to infer a particular phenotype (Table 3 and Supplementary Table 4).

### Genotype-phenotype correlation

Genotypes were classified according to the functional protein impact of the pathogenic *RPE65* variant (truncating or missense) and to the variant location at or out of the EL including the MBR of the RPE65 protein (Figure 1C). AAO of symptomatic disease, NB, VF reduction and VA loss distribution among variant class and location are detailed in Table 4.

**Table 4.**
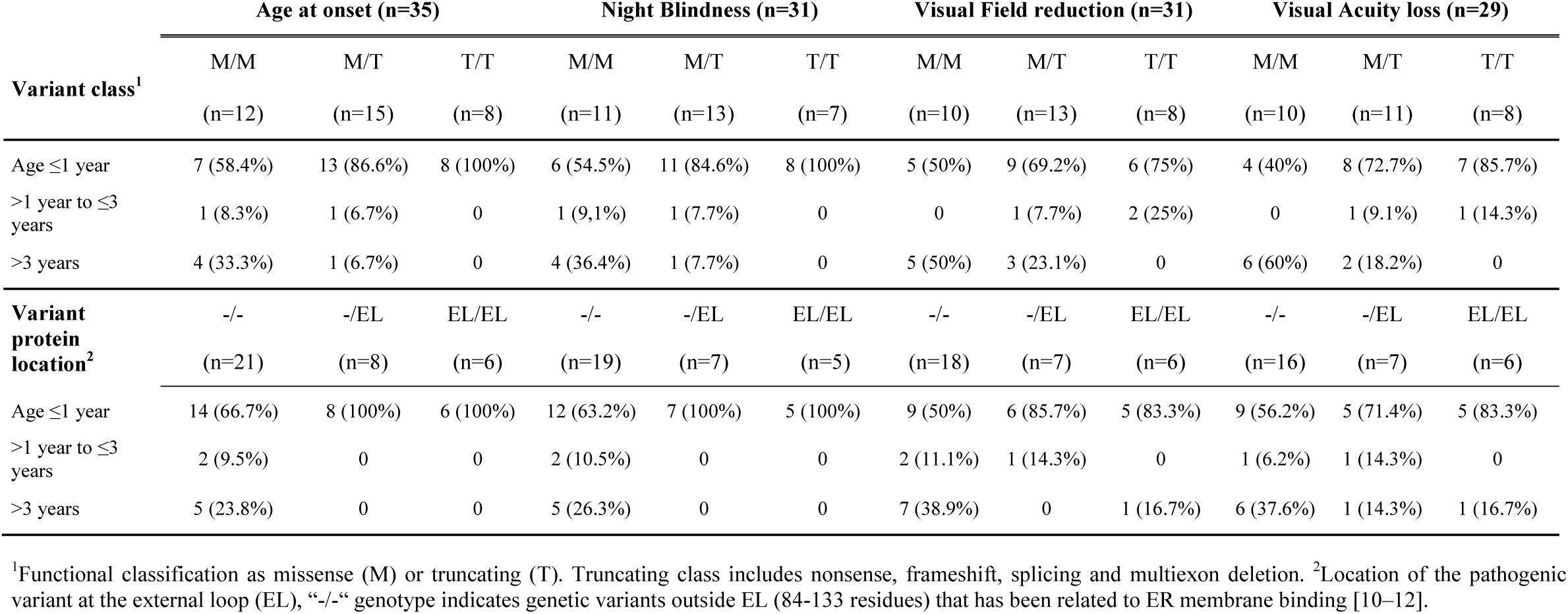
Age at onset of disease, night blindness, visual field reduction and visual acuity loss among patients according to the carried variant class and variant protein location.

| Variant class<sup>1</sup> | Age at onset (n=35) |  |  | Night Blindness (n=31) |  |  | Visual Field reduction (n=31) |  |  | Visual Acuity loss (n=29) |  |  |
| --- | --- | --- | --- | --- | --- | --- | --- | --- | --- | --- | --- | --- |
|  | M/M | M/T | T/T | M/M | M/T | T/T | M/M | M/T | T/T | M/M | M/T | T/T |
|  | (n=12) | (n=15) | (n=8) | (n=11) | (n=13) | (n=7) | (n=10) | (n=13) | (n=8) | (n=10) | (n=11) | (n=8) |
| Age ≤1 year | 7 (58.4%) | 13 (86.6%) | 8 (100%) | 6 (54.5%) | 11 (84.6%) | 8 (100%) | 5 (50%) | 9 (69.2%) | 6 (75%) | 4 (40%) | 8 (72.7%) | 7 (85.7%) |
| >1 year to ≤3 years | 1 (8.3%) | 1 (6.7%) | 0 | 1 (9.1%) | 1 (7.7%) | 0 | 0 | 1 (7.7%) | 2 (25%) | 0 | 1 (9.1%) | 1 (14.3%) |
| >3 years | 4 (33.3%) | 1 (6.7%) | 0 | 4 (36.4%) | 1 (7.7%) | 0 | 5 (50%) | 3 (23.1%) | 0 | 6 (60%) | 2 (18.2%) | 0 |
| Variant protein location<sup>2</sup> | -/- | -/EL | EL/EL | -/- | -/EL | EL/EL | -/- | -/EL | EL/EL | -/- | -/EL | EL/EL |
|  | (n=21) | (n=8) | (n=6) | (n=19) | (n=7) | (n=5) | (n=18) | (n=7) | (n=6) | (n=16) | (n=7) | (n=6) |
| Age ≤1 year | 14 (66.7%) | 8 (100%) | 6 (100%) | 12 (63.2%) | 7 (100%) | 5 (100%) | 9 (50%) | 6 (85.7%) | 5 (83.3%) | 9 (56.2%) | 5 (71.4%) | 5 (83.3%) |
| >1 year to ≤3 years | 2 (9.5%) | 0 | 0 | 2 (10.5%) | 0 | 0 | 2 (11.1%) | 1 (14.3%) | 0 | 1 (6.2%) | 1 (14.3%) | 0 |
| >3 years | 5 (23.8%) | 0 | 0 | 5 (26.3%) | 0 | 0 | 7 (38.9%) | 0 | 1 (16.7%) | 6 (37.6%) | 1 (14.3%) | 1 (16.7%) |

<sup>1</sup>Functional classification as missense (M) or truncating (T). Truncating class includes nonsense, frameshift, splicing and multiexon deletion. <sup>2</sup>Location of the pathogenic variant at the external loop (EL), “-/-” genotype indicates genetic variants outside EL (84-133 residues) that has been related to ER membrane binding [10–12].

The potential correlation among the variant type classified into 3 genotype groups and the AAO of first disease symptoms was studied. This analysis revealed that affected cases carrying 2 missense alleles presented significantly a later onset of the disease than those with 1 or 2 truncating variants (Log Rank test p=0.02; Figure 2C).

Similar findings resulted when patients were classified into 2 genotypes groups (missense/missense *vs*. truncating carrier, Log Rank test p<0.01). While near to 60% (7/12) of patients carrying a missense/missense genotype had an AAO ≤1 year, almost all patients (91%, 21/23) carrying at least 1 truncating allele presented symptoms before or at the first year of life (Table 4). A statistically significant association was also obtained for the AAO for the VA loss (p=0.04). A similar trend was observed for the AAO of NB and VF reduction (p=0.16 and 0.12, respectively), but did not reach statistical significance (Figure 2E-F). Our findings suggest that patients carrying 1 or 2 truncating alleles showed an earlier visual impairment compared to those carrying 2 missense variants (Table 4 and Figure 2). However, this should be taken with caution given the low number of individuals in each category (missense/missense= 12, missense/truncating= 15 and truncating/truncating= 8). In addition, affected subjects carrying variants located in the EL of the protein (Figure 1C) showed an earlier onset of the disease compared to those with 2 alleles outside this gene region (Table 4). In fact, all patients with at least 1 allele at the external loop region (n=14) showed an age at onset ≤1 year compared to the 66.7% of the patients without a pathogenic allele at that region (14 of 21), despite not being statistically significant (p=0.10; Supplementary Figure 1).

## DISCUSSION

Pathogenic variations in *RPE65* are causative of different IRD such as LCA, EORP or cone-rod dystrophies with AR pattern of heritance. *RPE65* variants explained the 2.6% of the AR non-syndromic IRD families genetically characterized in our 1042 families cohort [22], similar to the incidence (2.1%) found in previous studies of *RPE65*-associated IRD in European and North American populations [34] and higher than the prevalence of 0.8% recently reported in a Chinese cohort [19]. Besides, *RPE65* has been related to a considerable percentage (3-16%) of LCA cases worldwide [14–16, 19], in accordance to the near to 16% of *RPE65*-associated LCA found in our cohort. As LCA is the earliest and most severe form of IRD [2, 3], the better knowledge of *RPE65* pathogenic variants and their phenotypic spectrum is particularly relevant owing to the severity of the disease, the available therapeutic intervention and the ongoing clinical trials [24].

A total of 212 pathogenic variants in *RPE65* have been reported to date (HGMD, [17]), of which the majority are missense (54.71%), followed by nonsense-frameshift (32.54%) and splice site variants (12%). A similar frequency of the type of mutation was observed in our cohort (59%, 28% and 11%, respectively); thus, our cohort is representative of the *RPE65* prevalence and mutational spectrum. From the total of reported pathogenic variants, various alleles showed a high frequency in particular ethnicities such as p.(Tyr368His) in Dutch, p.(Glu102^*^) in Greek and p.(Arg91Trp) in Saudi Arabic populations, which may be related to putative founder effects [41]. In addition, 3 variants (p.(Arg91Trp), p.(Tyr368His) and c.11+5G>A) accounted for the 26.7% of all *RPE65* variants reported until 2016, as detailed by *Astuti et al* [41]. Remarkably, these common pathogenic *RPE65* alleles are not overrepresented in our cohort. Only 2 (p.Tyr368His and c.11+5G>A) of them were detected in our patients and with a lower frequency (6% and 4%, respectively). By contrast, the most common alleles found in our cohort were p.(Ile98Hisfs^*^26, p.(Pro111Ser) and p.(Gly187Glu). The p.(Ile98Hisfs^*^26) variant was first described in LCA patients from India [15] and Central America [42], being related to more severe phenotypes. Interestingly, p.(Ile98Hisfs^*^26) seems to represent a founder mutation in our cohort with all the carriers families sharing a common haplotype. Due to the historical influence of the Spanish ancestry in Central and South America populations, this finding could be related to the enrichment of this rare variant in the Latino cohort of *GnomAD* and to its increased frequency in LCA patients from Costa Rica where this variant was reported as a recurrent mutation [42]. The variant p.(Pro111Ser), which is carried by 3 of our families, were first described in our cohort of patients, as recently we reported [24]. This is an extremely rare variant for which only one allele was found in an African genome from the *GnomAD* database. The geographical origin of these unrelated families is Central and North Spain. Haplotype analysis revealed same alleles for 2 of the analyzed markers (D1S2803 and D1S448) and therefore, it is very likely that this variant arose from a founder event.

The 27 probands here reported were clinically diagnosed during the first months/years of life, with signs of congenital nystagmus, NB and VF progressive reduction and loss of central vision. In most of cases, ERG was completely extinguished as reported in literature [34, 39, 41]. Other ophthalmological symptoms such as severely disturbed colour vision in *RPE65*-associated LCA were also present in our cohort, in accordance with a previous report [1]. Interestingly, a patient from our cohort carrying the missense variants p.(Leu341Ser) and p.(Arg515Trp) displayed *fundus albipunctatus*-like changes, with congenital signs of impaired vision but BCVA values >0.3 in both eyes at the age of 10 years. Recently, in a cohort of 18 Chinese families with *RPE65*-associated IRD, a *fundus albipunctatus*-like phenotype has been described in 2 families also carrying the p.(Arg515Trp) variant in compound heterozygosis with other missense mutations, those patients retained good visual acuity and preserved photoreceptors along the years [19]. Additionally, the p.(Arg515Trp) in combination with the p.(Gln228Pro) was also found in a Japanese patient that displayed a *fundus albipunctatus*-like phenotype [43]. Therefore, these findings may indicate that the p.(Arg515Trp) variant could be specifically related to *fundus albipunctatus*-like phenotypes that rarely appears associated with other *RPE65* variants and thus, this variant may be causative of a milder *RPE65* phenotype.

Hypomorphic alleles in *RPE65* have been associated with milder forms of retinal dystrophies [44–46]. This fact suggests that those alleles would present lower levels of isomerase activity that may be enough to maintain an acceptable foveal cone function, despite the impaired rod and cone function reported by the anormal ERG records and a marked macular dysfunction in adulthood. Various hypomorphic alleles have been described along the *RPE65* gene, such as p.(Leu43Pro), p.(Arg91Trp), p.(Ala145Thr), p.(Tyr249Cys) and p.(Arg515Trp), all of them missense variants [44–46]. In our cohort, only p.(Arg515Trp) was detected in compound heterozygosis which phenotype was discussed above.

Symptomatic onset has also been associated with visual impairment severity. In fact, central VA loss seems to be more severe in adulthood when onset is before the first year of life compared to those patients with symptomatic beginning between one and five years of age [2, 18]. In this sense, the variant class may be related to symptomatic onset and could give some clues of the progression and severity of the associated phenotype. Our analysis reflected that missense variants were related to a later symptomatic onset of the disease in our cohort. According to this results, previous studies suggested that more severe phenotypes are caused by null mutations in both *RPE65* alleles, whereas milder forms result in cases of having at least one missense allele [14].

Missense variants may change the RPE65 protein structure by reducing or abolishing the enzyme activity or by changing its location [47]. Thus, not only the variant class but also its location at the protein could be relevant for disease phenotype. In our cohort, mutations are widely distributed across the *RPE65* gene but the most mutated region, with 5 pathogenic variants found in 10 of the 27 families was the external loop of the protein, which includes the ER membrane affinity residues and the anchor region [10, 11]. Our results pointed out to an early onset of the disease in those patients carrying mutations in this region despite of the kind of the variant, although this observation did not reach statistical significance. It is clear that RPE65 interaction with the ER membrane is critical for its enzymatic activity and the palmitoylated Cys112, in addition to the interaction with residues of the loop, may play an important role in the enzyme membrane affinity [11, 48]. Furthermore, the co-location of RPE65 and LRAT proteins is essential for their activity and alterations in their location contribute to decrease the enzymatic activities and their stability [48], affecting the visual cycle.

Retrospective design studies have limitations, thus statistical results should be interpreted with caution. In the present study, part of the data collection was based on historical notes from the ophthalmological clinical records, which impacts on data standardization, as some phenotypic information was incomplete and thus, limiting a deeper formal statistical analysis of genotype/phenotype associations. However, our design is appropriate for this cohort due to rarity incidence and the clinical characteristics of the disease. The limited information about phenotype correlations within the clinical spectrum of *RPE65* highlights the need for genetic testing. At the moment, genetic characterization becomes a key step not only in diagnosis but also in IRDs prognosis given that patients can benefit from different therapies. Gene therapy for the treatment of patients with confirmed biallelic pathogenic *RPE65* variants is approved [20, 21, 39]. Thus, it is particularly important to obtain information about the natural course of genotype-specific phenotypes for purposes of comparison with evolution before/after treatment.

In conclusion, our findings suggest an association between the functional impact of the *RPE65* variant carried and the age at onset of the disease, showing all patients carrying a truncating allele a symptomatic onset of the disease at the first year of life. Thus, our results provide useful information of *RPE65*-associated IRD phenotypes which may help to improve clinical and therapeutic management of these patients.

## Data Availability

All data referred to in the manuscript are included in the tables or supplementary materials. Any additional data will be available upon request to the corresponding author.

## ACKNOWLEDGEMENTS

We acknowledge the generous support from all patients and their families who participated in the study. We acknowledge Drs. Sorina Tatu and Raquel Perez-Carro for their help in genetic screenings, Dr. Belén Gil-Fournier from Getafe University Hospital, Drs. Guitart and Baena from Hospital Sabadell, Dr. Juan de Dios Garc^í^a from University Hospital Principe de Asturias, Madrid, Spain; Dr. Maria Isabel López-Molina from University Hospital Fundacion Jimenez Diaz, Dr. Leyre Juaristi from Donostia University Hospital, Dr. Loreto Martorell from Hospital Sant Joan de Deu, Dr. Ana Isabel Pastor Vivas from University Hospital Puerta de Hierro, for recruitment of patients and clinical data. We thank Ignacio Mahillo for the statistical support.

