## Supplementary material for "*RPE65*-related retinal dystrophy: mutational and phenotypic spectrum in 45 affected patients": ST1

Supplementary Table 1. List of primers used for confirming *RPE65* variants by Sanger sequencing.

| **Intron-Exon** | **cDNA (NM_000329.3)** | **Protein change (NP_000320)** | **Primer** |
| --- | --- | --- | --- |
| **IVS 1** | c.11+5G>A | p.? | F: gctcccaaagccataactc  R: aatcaatgccttctcttcagg |
| **IVS 2** | c.95-2A>T | p.? | F: gcccaatcaggctgctg  R: aagtgggtatataggttgcctcc |
| **Exon 2** | c.94G>C | p.Gly32Arg | F: ccagccctagagtgccttc  R: cctctccctgtgacccac |
| **Exon 3** | c.131G>A | p.Arg44Gln | F: gcccaatcaggctgctg  R: aagtgggtatataggttgcctcc |
|  | c.143G>A | p.Gly48Glu | F: gcccaatcaggctgctg  R: aagtgggtatataggttgcctcc |
| **Exon 4** | c.272G>A | p.Arg91Gln | F: ccctttattcttcatgttgtgc R: atttggagcttggaatggtc |
|  | c.292_311del | p.Ile98Hisfs*26 | F: ccctttattcttcatgttgtgc R: atttggagcttggaatggtc |
|  | c.311G>T | p.Gly104Val | F: ccctttattcttcatgttgtgc R: atttggagcttggaatggtc |
|  | c.331C>T | p.Pro111Ser | F: ccctttattcttcatgttgtgc R: atttggagcttggaatggtc |
| **Exon 5** | c.370C>T | p.Arg124* | F: ccctttattcttcatgttgtgc R: atttggagcttggaatggtc |
|  | c.457A>G | p.Thr153Ala | F: ccctttattcttcatgttgtgc R: atttggagcttggaatggtc |
| **IVS 5** | c.496-1G>A; | p.? | F: ttcaaggggtagtgatgacc  R: gagtaatttaactatgcacaaaatgc |
| **Exon 6** | c.514_515delGT | p.Val172Leufs*12 | F: ctctcaactggaggacattc R: tcagaagaggacagattggt |
|  | c.560G>A; | p.Gly187Glu | F: ctctcaactggaggacattc R: tcagaagaggacagattggt |
| **Exon 7** | c.722A>G | p.His241Arg | F:ctgctatttggattttcctg R:tcttcagaatcacaaacttg |
| **Exon9** | c.859delG | p.Val287fs*38 | F: catgtaggcactgttgattcttg R: ctcttgctgttttagatgtgattc |
|  | c.917C>T | p.Thr306Ile | F: catgtaggcactgttgattcttg R:ctcttgctgttttagatgtgattc |
| **Exon10** | c.1001_1002delTT | p.Phe334* | F: atggctctgatacacctggc R:gcttttgctaagtcacagtactcttc |
|  | c.1022T>C | p.Leu341Ser | F:atggctctgatacacctggc R:gcttttgctaagtcacagtactcttc |
|  | c.1102T>C | p.Tyr368His | F:atggctctgatacacctggc R:gcttttgctaagtcacagtactcttc |
| **Exon11** | c.1205G>A | p.Trp402* | F: tagcttcctgcagttcctcc  R: tctgatgggtatgaatcaggc |
| **Exon13** | c.1339C>T | p.Leu447Phe | F:cacacgggagtgaacaaatg R:aaggatcgtttttgagtattacgg |
|  | c.1341_1342dupCT | p.Cys448Serfs*4 | F: cacacgggagtgaacaaatg R: aaggatcgtttttgagtattacgg |
|  | c.1398C>G | p.Tyr466* | F:cacacgggagtgaacaaatg R:aaggatcgtttttgagtattacgg |
|  | c.[1388C>A; 1390_1410dup] | p.[Pro463His;Asp464_Pro470dup] | F:cacacgggagtgaacaaatg R:aaggatcgtttttgagtattacgg |
| **Exon14** | c.1451G>A | p.Gly484Asp | F: tcaggtcatatggttttctatatttg R: ggcctgtctcacagaggaag |
|  | c.1464T>A | p.Ser488Arg | F: tcaggtcatatggttttctatatttg R: ggcctgtctcacagaggaag |
|  | c.1543C>T | p.Arg515Trp | F: tcaggtcatatggttttctatatttg R: ggcctgtctcacagaggaag |
| **Exon1-14** | Chr1:68428822-68450322 del | p.? | *Multiplex ligation-dependent probe amplification* (Commercial kit, *MLPA, SALSA P221 LCA mix-1, MRC, Holland*) |
