## Supplementary material for "*RPE65*-related retinal dystrophy: mutational and phenotypic spectrum in 45 affected patients": ST2

Supplementary Table 2. List of microsatellites markers used to determine haplotypes of the *RPE65* locus.

| **Microsatellite marker** | **Start** | **End** | **Band** | **Primer Forward** | **Primer Reverse** | **Distance from *RPE65* (Mb)** |
| --- | --- | --- | --- | --- | --- | --- |
| D1S2806 | 67868153 | 67868355 | 1p31.3 | CATTACATCACAGCCTGATTAGA | CCACCATGCCTGACCT | 1,03 |
| D1S368 | 68231503 | 68231952 | 1p31.3 | GGGCATTGTTTAGGGGTG | TAGTGGGCTTTACGTCTGC | 0,66 |
| D1S2829 | 68252505 | 68252812 | 1p31.3 | AGTGGTTTATATGACTTACTGTGGG | GCACNCCAGCCTAGGTA | 0,64 |
| D1S2803 | 68918184 | 68918480 | 1p31.2 | AAATAAGTTCAAATCACAATCAGA | ATCTAACTCTGGGACTGGTAAA | intragenic |
| D1S448 | 69171301 | 68918480 | 1p31.2 | TGCAGAGATAGACTTTCGCT | AGTTAGTCATCCTTACCCAGC | 0,26 |
| D1S1162 | 69446498 | 69446935 | 1p31.2 | CCACACTATCATTTACCAGA | GGTTTCCTATGTTCCAAGC | 0,53 |
