## Supplementary material for "*RPE65*-related retinal dystrophy: mutational and phenotypic spectrum in 45 affected patients": ST3

**Supplementary Table 3.** *In silico* predictors of the novel RPE65 variants.

| Variant | SIFT Score | Polyphen-2 Score | MutationTaster Score | M-CAP Score | Other pathogenic predictors^1^ |
| --- | --- | --- | --- | --- | --- |
| Ex2:c.94G>C; p.Gly32Arg | Damaging | Probably damaging | Disease causing | Possibly Pathogenic | BayesDel_addAF, DANN, DEOGEN2, EIGEN, FATHMM-MKL, LIST-S2, MVP, MutationAssessor, PrimateAI, REVEL, and scSNV-Splicing |
| Ex3: c.143G>A; p.Gly48Glu | Damaging | Probably damaging | Disease causing | Possibly Pathogenic | BayesDel_addAF, DANN, DEOGEN2, EIGEN, FATHMM-MKL, LIST-S2, MVP, MutationAssessor, PrimateAI and REVEL |
| Ex6:c.514_515delGT; p.Val172Leufs*12 | NA | NA | NA | NA | GERP |
| Ex10: c.1001_1002delTT; p.Phe334* | NA | NA | NA | NA | GERP |
| Ex13: c.1339C>T; p.Leu447Phe | Damaging | Probably damaging | Disease causing | Possibly Pathogenic | BayesDel_addAF, DANN, DEOGEN2, EIGEN, FATHMM-MKL, LIST-S2, MVP, MutationAssessoR and REVEL |
| Ex13:c.1388C>A; p.Pro463His | Damaging | Probably damaging | Disease causing | Possibly Pathogenic | BayesDel_addAF, DANN, DEOGEN2, EIGEN, FATHMM-MKL, LIST-S2, MVP, MutationAssessor and REVEL |
| Ex13:c.1390_1410dup;p.Asp464_Pro470dup | NA | NA | NA | NA | GERP |
| Splice Variant^2^ | ***MaxEntScan***  ***[0-16]*** | ***Human Splicing Finder***  ***[0-100]*** | ***Splice Site Finder-like***  ***[0-100]*** | ***NNSPLICE [0-1]*** | ***GeneSplicer[0-15]*** |
| IVS5:c.496-1G>A;p.? | 9.2 → 0 | 85.4 → 76.8 | 84.2 → 74.2 | 0.98 → 0 | 7.1 → 0 |

^1^Information source:*VarSome: The Human Genomic Variant Search Engine*. Christos Kopanos, Vasilis Tsiolkas, Alexandros Kouris, Charles E. Chapple, Monica Albarca Aguilera, Richard Meyer, and Andreas Massouras. *Oxford Bioinformatics*, bty897, 30 October 2018. doi: <https://doi.org/10.1093/bioinformatics/bty897>

^2^Information source: *Alamut* software (*Interactive Biosoftware, Rouen, France*)
