## Supplementary material for "*RPE65*-related retinal dystrophy: mutational and phenotypic spectrum in 45 affected patients": ST4

**Supplementary Table 4.** Summary of clinical data in 45 affected subjects.

| **Family_ID** | **Patient** | **AAO of DS (years)** | **VF degree**  **(Age in yrs)** | **BCVA**  **(Age in yrs)** | **ERG**  **(Age in yrs)** | **OCT**  **(mac thick,**  **Age in yrs)** | **Cat**  **(yrs)** | **Other ocular pathology** | **Other remarks** |
| --- | --- | --- | --- | --- | --- | --- | --- | --- | --- |
| **RPE65-1** | P | <1yr | UNK | BE: 0.1 (<10yrs) | Alt (<5yrs) | UNK | N | Hyperopia and astigmatism | Developmental delay |
| **RPE65-2** | P | ≤1yr | UNK | BE: 0.16 (<20yrs) | Ext | UNK | N | Dyschromatopsia |  |
| **RPE65-3** | P | UNK | UNK | UNK | UNK | UNK | UNK |  |  |
| **RPE65-4** | P | <1yr | UNK | UNK | Ext | UNK | N |  |  |
| **RPE65-5** | P | ≤1yr | Non-specific central VF loss (<20yrs) | BE: 0.9 (<20yrs) | UNK | UNK | UNK | Myopia and astigmatism |  |
|  | S | ≤1yr | Non-specific central VF loss (30s) | RE: 0.7  LE: 0.9 (30s) | UNK | UNK | N | Myopia and astigmatism |  |
| **RPE65-6** | P | <20yrs* | UNK | RE: 0.01  LE: <0.01 (40s) | Alt (40s) | UNK | Y (40s) |  |  |
|  | S | <1yr | UNK | BE: 0 (30s) | Ext (30s) | UNK | UNK |  |  |
|  | S | <1yr | UNK | BE: 0.05 (<10yrs) | UNK | UNK | N |  |  |
|  | S | UNK | UNK | UNK | UNK | UNK | UNK |  |  |
| **RPE65-7** | P | <1yr | UNK | BE: 0.2 (<20yrs) | Ext | UNK | N |  |  |
|  | S | UNK | UNK | BE: 0.05 (20s) | Ext | UNK | UNK |  |  |
|  | S | UNK | UNK | RE: 0.1  LE: 0.05 (<20yrs) | Ext | UNK | UNK |  |  |
| **RPE65-8** | P | UNK | UNK | UNK | UNK | UNK | UNK |  |  |
|  | S | UNK | UNK | UNK | UNK | UNK | UNK |  |  |
|  | S | UNK | UNK | UNK | UNK | UNK | UNK |  |  |
| **RPE65-9** | P | <1yr | UNK | BE: perceive and project light (30s) | UNK | UNK | N |  | Obesity |
|  | S | <1yr | <10° (40s) | BE: 0 (40s) | UNK | UNK | UNK |  |  |
|  | S | <1yr | UNK | UNK | UNK | UNK | Y (20s) |  |  |
| **RPE65-10** | P | <1yr | 15° (<5yrs) | BE: 0.1 (<5yrs) | Alt | UNK | N | Myopia and dyschromatopsia | Mild psychomotor delay in the 1rst months of life |
| **RPE65-11** | P | <1yr | UNK | RE: 0.04  LE: 0.08 (<5yrs) | Ext (<5yrs) | UNK | N |  | Mild hearing loss (right ear) |
|  | S | <1yr | UNK | UNK | UNK | UNK | N |  |  |
| **RPE65-12** | P | ≤1yr | <10° (30s) | BE: <0.1 (30s); BE: HM (40s) | Ext | Complete atrophy of the outer layers affecting the fovea (40s) | UNK | Peripheral retinal telangiectasias and dyschromatopsia. Retinal detachment |  |
|  | S | ≤1yr | RE: 20°  LE: 10° (<10yrs) | RE: 0.1  LE: 0.05 (30s) | Ext | UNK | UNK | Myopia, astigmatism, dyschromatopsia |  |
| **RPE65-13** | P | ≤1yr | 10° central (<5yrs) | BE: 0.1 (<5yrs) | Alt | UNK | N | Myopia, astigmatism, dyschromatopsia | Developmental delay (mild language and psychomotor coordination affection) |
| **RPE65-14** | P | <1yr | Peripheral constriction (<5yrs) | BE: < 0.1 (<5yrs) | Ext | RE: 210µm  LE: 213µm (<10yrs) | N | Myopia, hyperopia, astigmatism and dyschromatopsia |  |
| **RPE65-15** | P | <1yr | UNK | BE: 0 (20s) | UNK | UNK | Y (40s) |  |  |
|  | S | UNK | UNK | UNK | UNK | UNK | UNK |  | Psychiatric disorder |
| **RPE65-16** | P | <1yr | 10° central (30s) | BE: 0.1 (20s) | Ext (20s) | RE: 205µm  LE: 202µm (30s) | N | Dyschromatopsia | Hydrocele, unaffected father with nystagmus |
| **RPE65-17** | P | ≤10yrs | BE: loss of superior and temporal fields (40s) | RE: CF 0.5m  LE: CF 1.5m (50s) | Alt (30s) | Reduced thickness (40s) | Y (50s) | Myopia Magna and dyschromatopsia |  |
|  | S | 20s | 10° central (50s) | BE: CF 2m (40s) | UNK | UNK | N |  | Dyslipidemia |
|  | S | ≤5yrs | UNK | RE: HM  LE: CF 2m (50s) | UNK | Macular athrophy, no edema (60s) | N | Myopia | Dyslipidemia |
|  | S | 20s | <10º central (50s) | LE: 0.2 (40s) | Ext (20s) | BE: 166µm. Macular atrophy, no edema (40s) | N | LE myopia, RE enucleated due to trauma |  |
| **RPE65-18** | P | ≤5yrs | Absolute Scotomae (30s) | BE: HM 0.5m (30s) | Ext (20s) | Total loss of photoreceptor line (30s) | Y (20s) | Photophobia |  |
| **RPE65-19** | P | ≤5yrs | 10° central (30s) | RE: 0.4  LE: 0.1 (30s) | UNK | RE: 215µm  LE: 209µm (30s) | Y (UNK) | Hyperopia and astigmatism; anterior uveitis in LE |  |
|  | S | UNK | 10° central (<20yrs) | RE: 0.4  LE: 0.3 (<20yrs) | UNK | BE: 187µm (<20yrs) | N | Bilateral hyperopia and astigmatism. N evidence of keratoconus |  |
| **RPE65-20** | P | ≤1yr | Non-specific central VF loss (<10yrs) | BE: 0.05 (<5yrs) | Alt | UNK | N | Astigmatism | Obesity |
| **RPE65-21** | P | <1yr | UNK | RE: 0.4  LE: 0.3 (<10yrs) | UNK | UNK | N | Astigmatism |  |
| **RPE65-22** | P | <1yr | UNK | BE: HM (20s) | UNK | One line in BE. Atrophy of outer layers, choroidal folds RE>LE (20s) | Y (<1yr) | Congenital glaucoma, and optic atrophy. |  |
|  | S | UNK | UNK | UNK | UNK | UNK | UNK |  |  |
| **RPE65-23** | P | <1yr | UNK | UNK | UNK | UNK | N |  |  |
| **RPE65-24** | P | <1yr | UNK | UNK | UNK | UNK | N |  |  |
| **RPE65-25** | P | <1yr | UNK | BE: 0.1 (<5yrs) | UNK | UNK | N |  |  |
| **RPE65-26** | P | <1yr | UNK | BE: 0.1 (<5yrs) | UNK | UNK | N | Hyperopia |  |
| **RPE65-27** | P | <1yr | <10º (30s) | BE: 0 (30s) | UNK | UNK | N | Hyperopia, astigmatism and strabismus (RE) |  |

Sx, Sex; AAO of DS, Age at onset of disease symptoms; VF, Visual Field; BCVA, best-corrected visual acuity in decimal scale; ERG, full-field electroretinography; OCT, optical coherence tomography; mac thick, macular thickness; Cat, Cataracts; P, Probandus; S, Sibling; yr, year; yrs, years; UNK, Unknown; BE, both eyes; RE, right eye; LE, left eye; HM, Hand motion; CF, Counts fingers; Alt, Altered; Ext, Extinguished.

*Indicates age at Dx, patient refers AAO at infancy.
