## Supplementary material for "*RPE65*-related retinal dystrophy: mutational and phenotypic spectrum in 45 affected patients": Sup Figure legend

**Supplementary Figures Legends.**

**Supplementary Figure 1.** Age at onset of disease among patients according to the carried variant location.

A) Log Rank test and survival curve (free of disease) of the AAO of the first sign of disease, B) NB, C) VF reduction and D) VA loss according to patients’ genotype (0, 1 or 2 alleles located at the external loop of the RPE65 protein).

AAO, Age at onset; NB, night blindness; VF, Visual Field; VA, Visual Accuity; ns, not significant; EL, external loop; -/-, carrier of two variants outside the EL; -/EL, carrier of one allele at the EL and EL/EL, carrier of two *RPE65* alleles at the EL.
