## Supplementary figures and images for "*RPE65*-related retinal dystrophy: mutational and phenotypic spectrum in 45 affected patients"

### Sup Figure 1

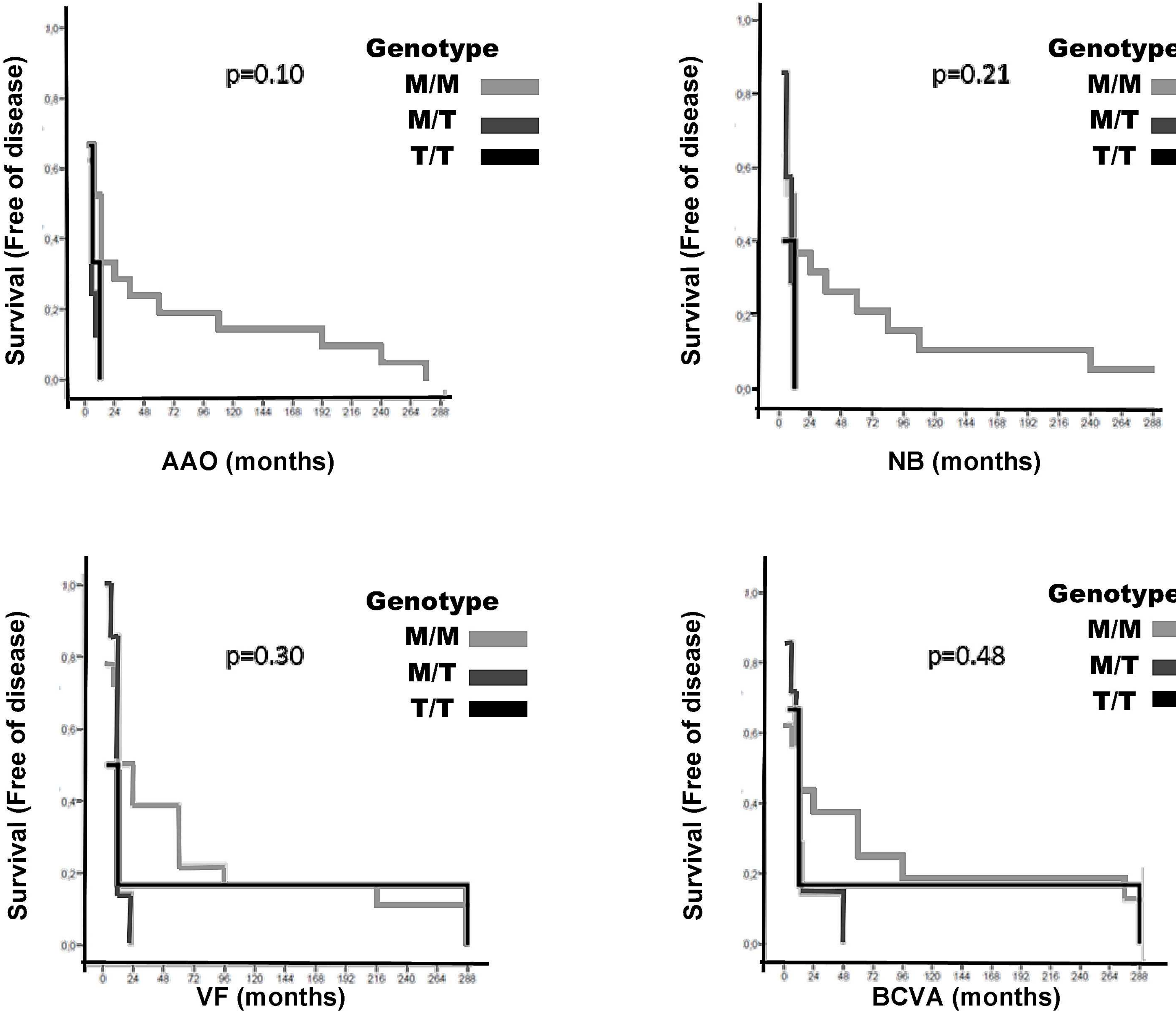
